# Association between trajectories of breastfeeding duration and infant health: Prospective cohort study

**DOI:** 10.64898/2026.11.02.26367067

**Authors:** Tatiana Dessy, Cindy-Lee Dennis

**Affiliations:** Lunenfeld-Tannenbaum Research Institute, Mount Sinai, Toronto, ON, Canada; Lawrence S Bloomberg Faculty of Nursing, Toronto, ON, Canada

**Author notes:** **Corresponding Author:** Tatiana Dessy, PhD, Lunenfeld-Tanenbaum Research Institute, 600 University Ave, Toronto, ON, Canada, M5G 1X5.

## Abstract

**Background:** Breastfeeding is a modifiable pathway contributing to infant health.

**Objectives:** To estimate trajectories of breastfeeding and the association between trajectories of breastfeeding and infant health.

**Methods:** We conducted a prospective longitudinal cohort study (2014-2020, Canada) with 3217 cohabitating English-literate mother-father-infant triads with internet or telephone access. Women with major psychiatric conditions or suicidality were excluded. Data were collected online at baseline (<3 weeks postpartum), 3, 6, 9, 12, 18, and 24 months. Breastfeeding was measured at 0-12 months. Baseline measures included attention-deficit disorder with/without hyperactivity (ADHD, Adult ADHD Self-Report Scale) and childhood adversity (Adverse Childhood Experiences International Questionnaire). We measured infant health at 12-24 months using the Brief Infant-Toddler Social and Emotional Assessment (challenging behaviors) and self-reported number of emergent visits (>90^th^ percentile). We estimated trajectories of breastfeeding using latent class growth analysis. Associations were estimated using adjusted linear mixed models.

**Results:** Most included mothers (n=2548) were of Canadian birth (2078/2548, 82%) and graduated from university (1920/2548, 75%); few reported unemployment (222/2548, 9%) or prenatal depression (240/2548, 9%). We identified two trajectories of breastfeeding: Constant Breastfeeding (1522/2548, 60%) and Decreasing Breastfeeding (1026/2548, 40%). No statistically significant differences were observed between classes for either the risk of challenging behaviors (RR −0.006 95% CI −0.028 to 0.016) or the risk of emergent visits (RR −0.015 95% CI −0.053 to 0.023).

**Conclusions:** In this study, discontinued breastfeeding was not associated with infant health. Future studies should focus on 6-month exclusive breastfeeding, a modifiable therapeutic infant feeding target.

**Article Summary:** In this prospective community-based cohort of 2548 Canadian mother-father-infant triads, we observed that maternal lifetime risk factors of depression – ADHD and childhood adversity – were associated with increased risk of infant morbidity from 12-24 months, but not decreasing vs. constant breastfeeding from 0-12 months. Breastfeeding duration was not significantly associated with infant morbidity.

## Introduction

Breastfeeding is a modifiable exposure predicting infant health [1]. The most robust evidence of a protective effect of breastfeeding on infant health has been observed in an experimental study of the impact of a breastfeeding promotion intervention on infant immunity [2]. In their 2001 study Kramer et al. observed higher rates of 6-month exclusive breastfeeding in the intervention group; a 40% reduction in gastrointestinal infections; and a 46% reduction in atopic eczema within 17046 mother-infant pairs (full-term and normal weight) at 12 months.

Maternal mental health is another well-established predictor of infant health: cumulative maternal distress – spanning pre-conception to perinatal periods – is correlated with increased risk of infant developmental challenges, including adverse birth outcomes, emotional dysregulation, and frequent emergent visits in the first years of life [3–6]. However the evidence supporting a causal effect of maternal mental health on infant health is less robust than for breastfeeding, by study design. One experimental study of a postpartum mental health promotion intervention on infant development found no effect on infant development [7].

In this prospective longitudinal cohort study including up to 3217 mother-father-infant triads from a Canadian community-based cohort, we aim to estimate trajectories of breastfeeding, the association between maternal lifetime risk factors of depression – ADHD and childhood adversity – and breastfeeding duration, as well as the association between these risk factors and infant morbidity at 12-24 months (socioemotional and physical health) [8]. We hypothesize that mothers will differ in breastfeeding duration due to underlying variability in mother, infant, and family psychosocial, mental, and physical health. We also emit the hypothesis that maternal ADHD and childhood adversity impact an infant’s development at 12-24 months through mother-child immune dysregulation pathways and breastfeeding.

## Methods

### Study Design and Participants

We conducted a 24-month prospective longitudinal cohort study using data from a Canadian community-based postnatal mental health cohort study (2014-2020) [8]. Participants were 3217 mother-father-infant triads recruited via social media, in-hospital, and through community outreach programs. Inclusion criteria were: having a recent singleton birth within 3 weeks postpartum at >33 weeks gestation; cohabitating biological parents; being English-literate; and having internet or telephone access. Owing to ethical concerns, women were excluded from the study if they had major psychiatric conditions (ie, schizophrenia, bipolar disorder, or psychosis) or were experiencing suicidality. All participants provided informed consent upon recruitment. This study was conducted in accordance with STROBE reporting guidelines.

### Ethical Approval

We conducted a secondary analysis based on data collected in a previous study which received ethical approval by the [*blinded*] Institutional Research Ethics Board (#29655) and participating health centers. Further ethical approval was not needed to conduct this study.

### Measures

#### Breastfeeding

The primary exposure was breastfeeding status from 0-12 months, assessed via maternal reports on infant feeding. Women were asked: ‘*Are you currently breastfeeding or giving your baby expressed breast milk?*’ at baseline, 9, and 12 months. At 3-6 months, women were asked: ‘*In the past 7 days, have you given your baby any breast milk?*’. Answering “yes” to either question indicated that the infant was breastfed.

#### Lifetime Risk Factors of Depression

Secondary exposures included parental ADHD and childhood adversity, two well-established risk factors of future depression or other mental health illnesses [9,10]. The psychometrically-validated ASRS (Part A) was used to assess symptoms of inattentiveness, hyperactivity, and impulsivity characterizing ADHD [11]. We calculated sum scores and applied the clinical cut-off ≥14 to identify mothers and fathers who screen positive for ADHD [12]. The psychometrically-validated ACE-IQ was used to assess 13 types of childhood adversity: neglect (emotional, physical); abuse (emotional, physical); violence (against a household member, in community, or collective); bullying; having a precarious family composition (single-parent household, parental separation/divorce, or being an orphan); and living with a household member who misused substance, was mentally ill/suicidal, or was incarcerated/jailed [13]. We calculated sum scores and applied clinically-meaningful cut-off >6 to identify childhood adversity.

#### Infant Health

The primary outcome was infant socio-emotional health from 12-24 months. We modelled challenging behaviors (socio-emotional health problems) using the 31-item psychometrically-validated BITSEA “problems” subscale completed by the mother at 12, 18, and 24 months [14]. We calculated sum scores and applied age and sex-specific clinical cut-offs to identify infants at higher risk of socio-emotional developmental delays and ASD [15]. The secondary outcome was infant physical health from 12-24 months. We calculated the 90^th^ percentile of the total number of emergent visits over the past six months reported by the mother at 18 and 24 months. We applied this cut-off to identify infants at higher risk of physical morbidity.

#### Covariates

We measured baseline socio-demographics for mothers and fathers, including: being Canadian born, having a university diploma, being unemployed. We measured substance use during pregnancy in mothers and fathers, including: alcohol, cigarettes, and other substances (unspecified). We assessed mental health during pregnancy via self-reported prenatal depression and anxiety in mothers and fathers. We measured the following infant characteristics using baseline maternal reports: sex assigned at birth, preterm birth, NICU use, and low birth weight. We also assessed ASD using the M-CHAT completed by the mother at 24 months [16]. We calculated sum scores and applied the clinical cut-off ≥3 to identify infants with probable ASD.

### Data Availability Statement

The raw and deidentified data used in this study are available upon reasonable request to the senior author for planned analyses, with permission from the research team.

### Statistical Analysis

Participants were described using absolute and relative frequencies or median and IQR. Reported characteristics included mother and father socio-demographics (including ADHD and childhood adversity), prenatal substance use and mental health, and mother-reported infant characteristics. Included participants had complete responses to breastfeeding and BITSEA items at all time points.

Trajectories of breastfeeding from 0-12 months were estimated via LCGA using R package lcmm [17]. A pre-specified number of three latent classes were retained provided they were interpretable. Non-linear probability models were fit with three equidistant splines as a link function. The model included a random intercept. Maximum likelihood estimation was used on complete-cases. The associations between trajectories of breastfeeding and lifetime risk factors of depression (ie, maternal ADHD and childhood adversity) were estimated using Cohen’s *d* with 95% CI.

We estimated linear mixed models of infant health outcomes measured from 12-24 months adjusted for trajectories of breastfeeding, time (0, 6, 12 months elapsed), maternal childhood adversity (number of ACE), and parental ADHD (sum scores) using R package lme4. Models were weighted using the posterior probabilities of the trajectories. We reported RR for each trajectory of breastfeeding. We calculated heteroskedasticity-robust SE if the random effects were significantly different from zero and derived 95% CI based on normality of the estimator. All mixed models included random effects for the intercept and time and a fixed quadratic term for time, if convergence criteria were satisfied. In sensitivity analyses, we used continuous measures of infant health.

Missing data were described using absolute and relative frequencies for the following variables: maternal ADHD and childhood adversity reported at baseline and the number of emergent visits reported at 18 and 24 months. If missingness was >5% and <40%, multiple imputation by chained equations with m= 10 iterations was used to improve precision.

All statistical analyses were conducted using R version 4.6.0 [18].

## Results

### Participant Characteristics

We included 2548mother-father-infant triads with complete data for breastfeeding from 0-12 months and BITSEA scores at 12-24 months. The majority of participants were Canadian born and few had experienced clinically significant childhood adversity. Most mothers were university educated and few were unemployed. The prevalence of ADHD was low and few used cigarettes, cannabis, or illegal substances during pregnancy, though nearly to one in five drank alcohol. Few self-reported prenatal depression; prenatal anxiety was more common (nearly one in five). Conversely, fewer fathers were university education and unemployed. Most used substances during pregnancy – especially alcohol – and frequencies of self-reported prenatal depression or anxiety were low. The prevalence of ADHD was higher in fathers compared to mothers (nearly one in ten). Infant health characteristics indicated that low birth weight and ASD were uncommon, though either preterm birth or NICU use was observed in one in ten infants.

### Primary Findings

#### Trajectories of Breastfeeding

We estimated two distinct trajectories of breastfeeding from 0-12 months (Figure 1). The estimated standard deviation for the random intercept was SD 1.26. Most participants were clustered in the “Constant Breastfeeding” class (n= 1522/2548, 60%) whereas the remaining were clustered in the “Decreasing Breastfeeding” class (n=1026/2548, 40%). The proportion of mothers with ADHD was <5% in the Constant and Decreasing Breastfeeding classes. A higher proportion of mothers with clinically significant childhood adversity were clustered in the Constant Breastfeeding class (n= 177/1522, 17%) compared to the Decreasing Breastfeeding class (n= 100/1026, 10%). The association between maternal lifetime risk factors of depression and trajectories of breastfeeding was not statistically significant: Cohen *d* 0.11, 95% CI −0.10 to 0.32 for ADHD; Cohen *d* 0.10, 95% CI −0.03 to 0.22.

**FIGURE 1.**
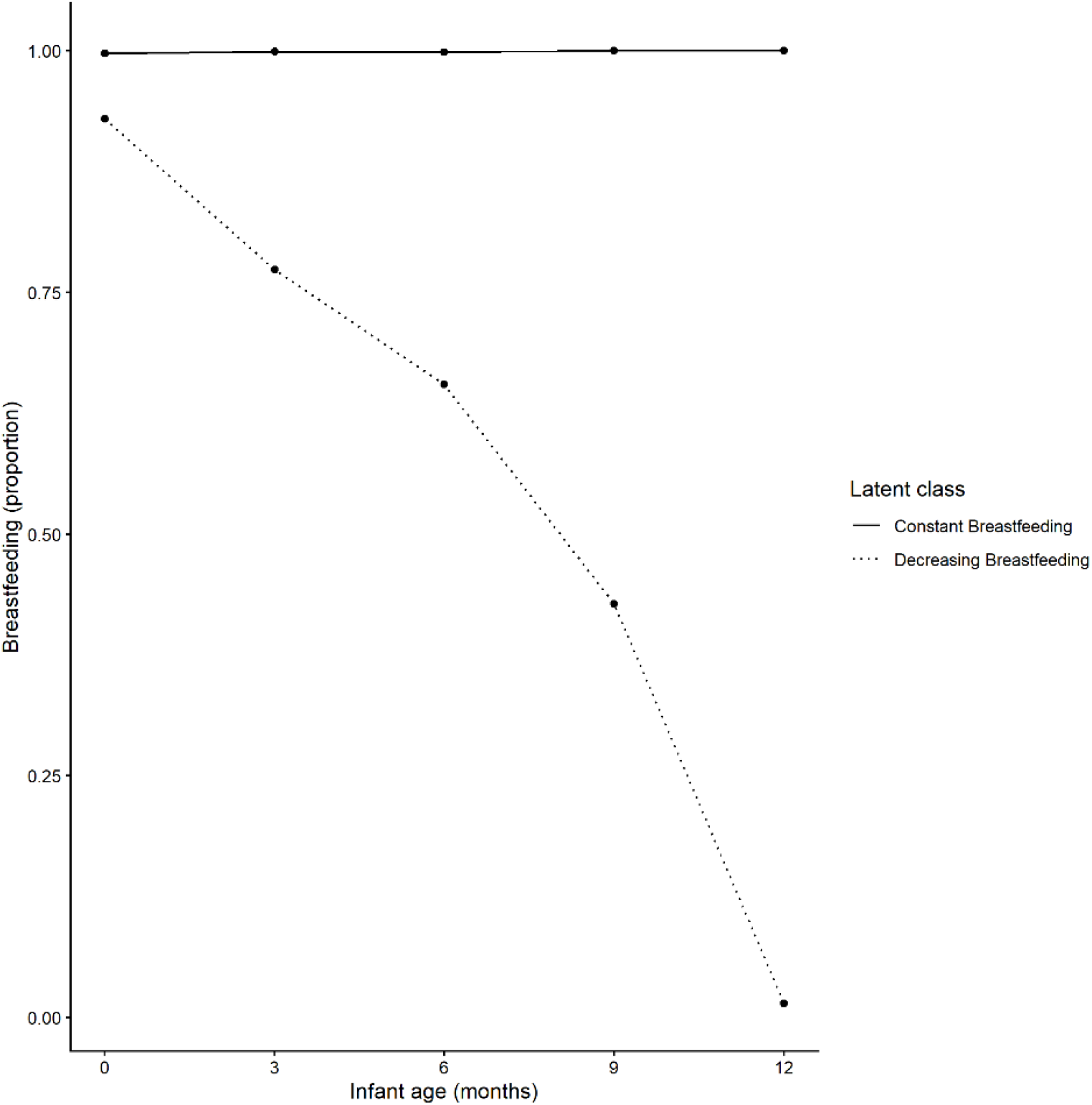
Trajectories of breastfeeding duration. Trajectories were estimated using Latent Class Growth Analysis with non-linear probability models fit using three equidistant splines. The proportions of participants in each class were: 1522/2548 (60%) in “Constant breastfeeding” and 1026/2548 (40%) in “Decreasing Breastfeeding”.

#### Infant Health

From 12-24 months, the proportion of infants with clinically significant challenging behaviors decreased in the Constant Breastfeeding cluster vs. increased in the Decreasing Breastfeeding cluster; the mean number of emergent visits decreased in both clusters (Figure 2-3). In models of infant health with categorical outcome specifications, there was no statistically significant association between Decreasing Breastfeeding and infant morbidity (Table 2). The estimated random intercept SD were 0.16 in the model of infant challenging behaviors and 0.17 in the model of infant emergent visits. However, higher maternal ADHD scores and higher number of maternal adverse childhood experiences were associated with increased risk of infant challenging behaviors: RR 0.011, 95% CI 0.007 to 0.014 for maternal ADHD and RR 0.011, 95% CI 0.006 to 0.016 for maternal childhood adversity, given paternal ADHD scores and within the Decreasing Breastfeeding class. Similar results were obtained in models using continuous outcome specifications. Missing data were <5% for parental ADHD and maternal childhood adversity, but >40% (n=1429/2548) for the number of emergent visits; therefore, multiple imputation was not used.

**TABLE 1.** Characteristics of included study participants (2014-2020, Canada).

|  | **No. (%)** |  |
|---|---|---|
|  | **Mothers**<br>**(N=2548)** | **Fathers**<br>**(N=2548)** |
| **Socio-demographics** |  |  |
| Canadian born | 2078 (82) | 1994 (78) |
| University diploma | 1920 (75) | 1433 (56) |
| Unemployment | 222 (9) | <sup>†</sup> |
| Childhood adversity, ACE-IQ | 277 (11) | 302 (12) |
| ADHD, ASRS | <sup>†</sup> | 203 (8) |
| **Prenatal Substance Use** |  |  |
| Alcohol | 403 (16) | 1986 (78) |
| Cigarettes | <sup>†</sup> | 273 (11) |
| Other substances | <sup>†</sup> | 179 (7) |
| **Prenatal Mental Health** |  |  |
| Depression | 240 (9) | 123 (5) |
| Anxiety | 372 (15) | 237 (9) |
| **Infant Characteristics** |  |  |
| Sex assigned at birth | 1300 (51) |  |
| Preterm birth or NICU use | 267 (11) |  |
| Low birth weight | <sup>†</sup> |  |
| ASD, M-CHAT, 24 months | <sup>†</sup> |  |

Abbreviations: ACE-IQ, Adverse Childhood Experiences International Questionnaire; ADHD, attention-deficit disorder with/without hyperactivity; ASD, autism spectrum disorders; ASRS, Adult ADHD Self-Report Scale; IQR, interquartile range; M-CHAT, Modified Checklist for Autism in Toddlers; NICU, neonatal intensive care unit.
<sup>†</sup>Cell frequency <5%.

**TABLE 2.** Association between trajectories of breastfeeding duration and infant health.

|  | **Breastfeeding, Decreasing vs Constant**,<br>**RR (95% CI)** |
|---|---|
| **Challenging behaviors, BITSEA** | -0.006 (-0.028 to 0.016) |
| **Physical morbidity, number of emergent visits >90<sup>th</sup> percentile** | -0.015 (-0.053 to 0.023) |

Abbreviations: BITSEA, Brief Infant-Toddler Social and Emotional Assessment; CI, confidence interval; RR, risk ratio; SD, standard deviation; SE, standard error.
Showing results from linear mixed models including a random intercept and fixed effects for latent class (Decreasing Breastfeeding vs. Constant Breastfeeding), parental ADHD, maternal childhood adversity, and time; using complete-cases only, with maximum likelihood estimation.  Note that 56% of observations on emergent visits at 18-24 months were missing from the analytic sample.

**FIGURE 2.**
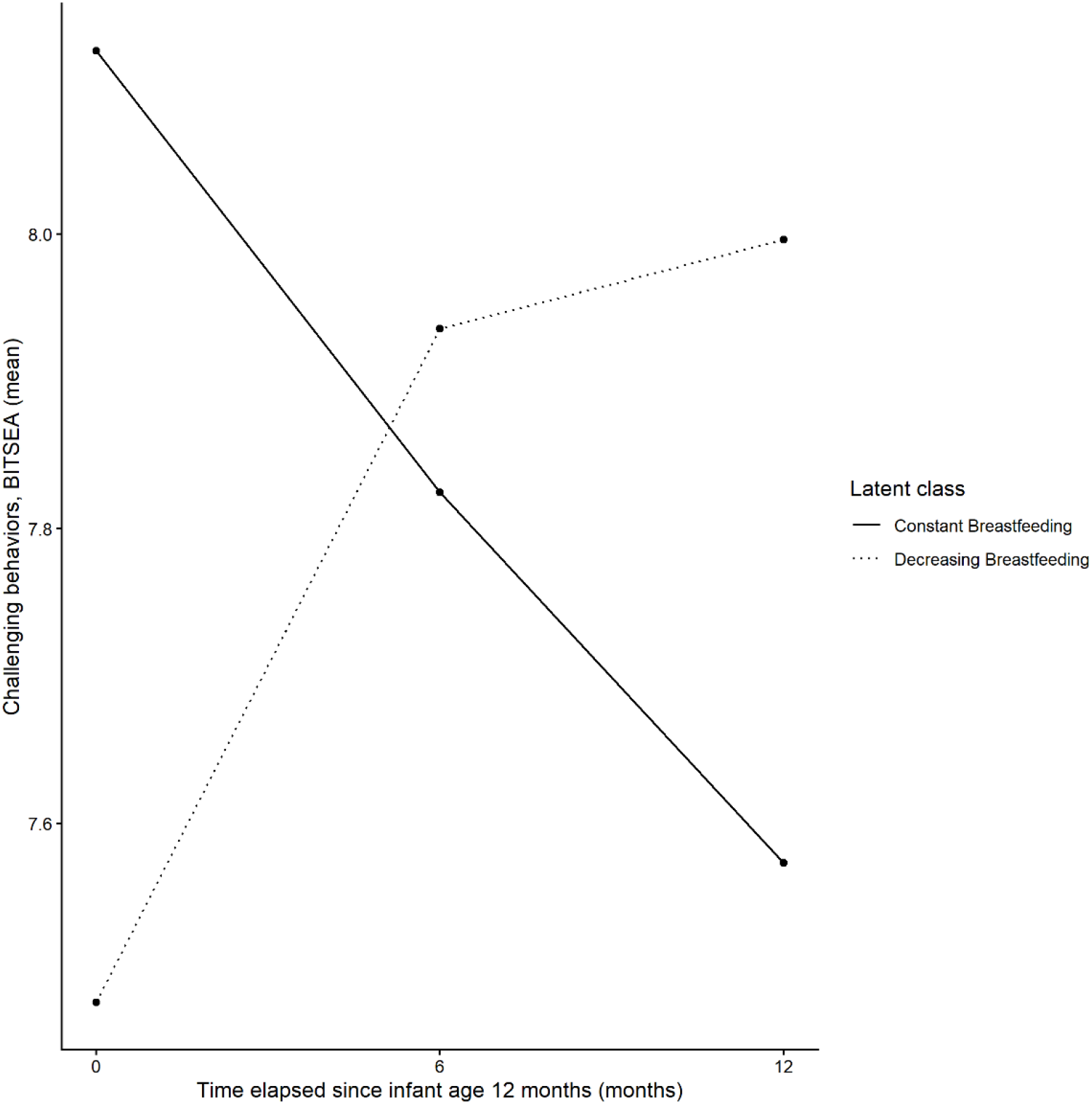
Infant challenging behaviors by trajectory of breastfeeding duration, across time. Abbreviations: BITSEA, Brief Infant-Toddler Social and Emotional Assessment.

**FIGURE 3.**
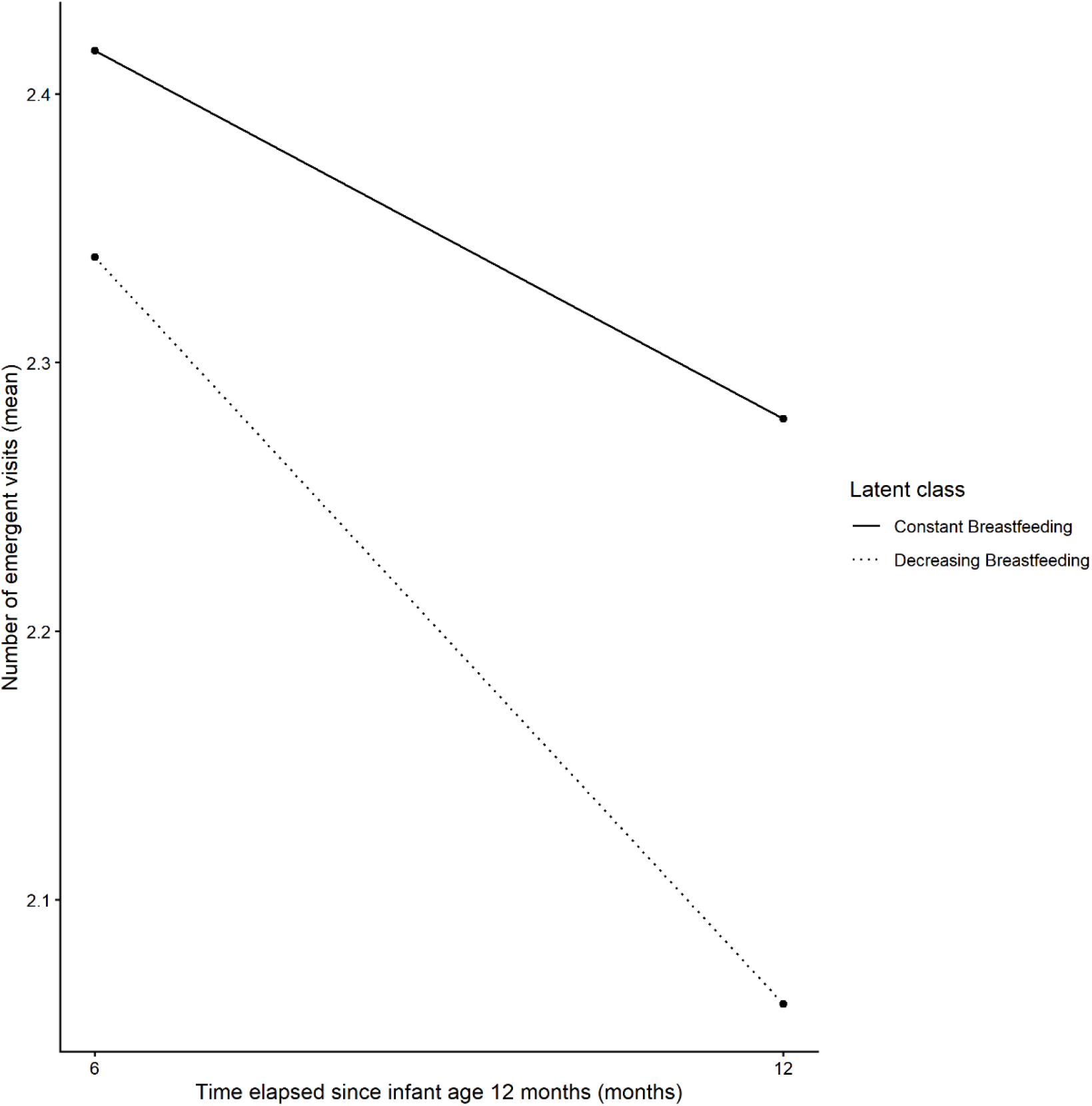
Infant emergent visits in the past 6 months by trajectory of breastfeeding duration, across time.

## Discussion

Overall, we observed no statistically significant association between trajectories of breastfeeding duration (Constant vs. Decreasing) from 0-12 months and infant health (socio-emotional or number of emergent visits) from 12-24 months. However we observed a statistically significant increase in challenging behaviors at 12-24 months with increasing maternal ADHD and ACE. Our study contributes to the existing literature by focusing on breastfeeding duration, as measured by Constant vs Decreasing breastfeeding classes, and its association with infant health, considering maternal lifetime risk factors of depression. Our findings indicate that breastfeeding of any kind (exclusive or mixed) is not a relevant predictor of infant health, unlike exclusive breastfeeding.

To our knowledge, this is the first study to examine the relationship between breastfeeding duration and infant socio-emotional health as measured by the BITSEA. However parental anxiety and depression were previously observed to be associated with poorer socio-emotional health in 1004 children >5 years of age, though infant socio-emotional health was measured using other scales [19].

The relationship between maternal lifetime risk factors of depression and infant health have been reviewed. Parental childhood adversity measured by ACE questionnaires was associated with an increasing number of emergent visits in early life in a meta-analysis of 15 studies [20]. In a separate study, maternal depression measured by Edinburgh Postnatal Depression Scale scores was observed to be associated with an increasing number of emergent visits in the first year of life [3].

Our findings may not be generalizable to those families with lower socioeconomic status and important psychiatric comorbidities owing to sampling limitations and study exclusion criteria.

### Strengths and Limitations

Our study’s strengths include the prospective longitudinal design, with data spanning 0-12 months for the exposure (breastfeeding duration) and 12-24 months for the outcome (infant health). Owing to our study design, the risk of confounding by indication and reverse causation is minimal. We also used psychometrically-validated questionnaires, where possible, to measure infant health and maternal lifetime risk factors of depression, which minimizes measurement errors.

One important limitation is that infant was measured after the usual etiological window (first year of life) [2,3]. We also had a significant number of missing observations for the number of emergent visits (>40%), which impeded the use of techniques such as multiple imputation to increase statistical precision. The reason for the emergent visit was unspecified, therefore we could not study specific causes or illnesses like previous studies [2]. Finally, the use of self-report questionnaires presents an increased risk of misclassification errors, which could bias our estimates towards the null.

## Conclusions

In this prospective community-based cohort of 2548 Canadian mother-father-infant triads, we examined the relative contributions of maternal lifetime risk factors of depression – ADHD and childhood adversity – and breastfeeding trajectories to infant socio-emotional and physical health. We observed that infant morbidity was not associated with decreased breastfeeding or maternal lifetime risk factors of depression. Our findings indicate that breastfeeding duration does not contribute to infant morbidity, as measured by challenging behaviors and emergent visits – for any reason – from 12-24 months. However mothers’ ADHD and childhood adversity may predict infant socio-emotional challenges, which could inform early identification and prevention for infants at higher risk of morbidity. Infant health remains better predicted by exclusive breastfeeding rather than duration of any breastfeeding.

## Abbreviations

ACE-IQ: Adverse Childhood Experiences International Questionnaire
ADHD: Attention-deficit disorder with/without hyperactivity
ASD: Autism spectrum disorders
ASRS: Adult
ADHD: Self-Report Scale
BITSEA: Brief Infant-Toddler Social and Emotional Assessment
IQR: Interquartile range
LCGA: Latent Class Growth Analysis
M-CHAT: Modified Checklist for Autism in Toddlers
NICU: Neonatal intensive care unit
RR: Risk ratio
SMD: Standardized mean difference

## Conflict of Interest Disclosures (includes financial disclosures)

The authors have no conflicts of interest relevant to this article to disclose.

## CONTRIBUTORS STATEMENT PAGE

### Contributors’ Statement

Dr Tatiana Dessy conceptualized and designed the study, cleaned the raw data, planned and conducted the statistical analyses, drafted the initial manuscript, and critically reviewed and revised the manuscript for important intellectual content.

Dr Cindy-Lee Dennis provided access to the IMPACT dataset and critically reviewed and revised the manuscript.

All authors approved the final manuscript as submitted and agree to be accountable for all aspects of the work.

